# Interpretable Machine Learning for Epileptic Seizure Detection on the BEED Using LIME with an Ensemble Network

**DOI:** 10.1101/2025.09.30.25336996

**Authors:** Biplov Paneru

## Abstract

This study aims to identify seizures in four different stages among epileptic patients, utilizing the Bangalore Epilepsy Dataset (BEED). This dataset, which has 16 channels, was sourced from the UCI Machine Learning Repository. Initially, the data underwent preprocessing through UMAP for dimensionality reduction. This was succeeded by feature extraction via the Fast Fourier Transform (FFT), which transformed the scaled signals into the frequency domain to capture their spectral characteristics. The findings show that a two-level ensemble model surpasses the performance of leading methods, reaching an accuracy rate of 97.06%. The model’s performance was confirmed through stringent nested cross-validation, guaranteeing consistency across all dataset folds. The model’s potential for real-time deployment on Edge and Internet of Things (IoT) devices is underscored by these findings.

## Introduction

As machine learning (ML) and artificial intelligence (AI) continue to advance, there is an increasing emphasis on using these technologies to improve clinical applications. Early disease diagnosis and prediction to facilitate early preventive intervention is a major objective in healthcare [1]. Researchers have focused a lot of emphasis on epileptic seizures, which are neurological disorders marked by particular patterns in electroencephalography (EEG) recordings. EEG signal feature extraction and pattern classification have been successfully accomplished using machine learning (ML) and deep learning (DL) approaches [2]. Recurrent seizures that result in unconsciousness or muscle spasms are a common neurological disorder called epilepsy, which has a major negative influence on a patient’s health, everyday activities, and general well-being [3].

Because EEG recordings offer vital information about the electrical activity of the brain, they are widely employed in epilepsy research [4]. EEG data analysis using ML and DL techniques has been the subject of numerous studies aimed at identifying and forecasting epileptic episodes [5, 10, 12]. More accurate seizure identification has been made possible by developments in EEG processing techniques, such as improved feature extraction methods, improved artifact removal, and improved time-frequency analysis [6]. About 50 million individuals worldwide suffer from epilepsy, which is one of the most common and dangerous neurological conditions, affecting 1% to 2% of the world’s population [7]. Given the severity of seizures and their risks, alongside the pressing demand for more accurate and advanced therapeutic approaches have been developed to identify seizure activity and deliver targeted electrical stimulation to help suppress seizures and reduce their frequency [9].

This work solely focuses on development of hybrid models as well as ensemble, traditional ML models to find out the potentiality for deployment for aiding patient with epileptic seizure along with classifying it. The proposed research is divided into various section in which section I. is related to abstract, and general introduction along with related works. Section 2. Refers to methodology section in which various ML models are proposed and architecture proposed are shown. Section 3. Shows the results from the models as well as discussions from the results obtained finally. Comparing work with previous works and discussion on limitations of study. Final section deals with concluding the study describing its outcomes and potential application for real-world epilepsy related effects minimization with AI-powered technology. The work provides a framework for utilizing applications-based on the early seizure detection for the aiding of patients at the emergency situations.

## Related works

Recent years have seen substantial advances in machine learning (ML) techniques for epileptic seizure prediction and detection using EEG signals. Rasheed et al. [1] provided a comprehensive review highlighting state-of-the-art ML methods for early seizure prediction, identifying research gaps, challenges, and suggesting future directions to enhance model robustness and generalizability.

Several works have employed the UCI Epileptic Seizure Recognition dataset for classification tasks. Kode et al. [2] evaluated multiple classifiers, including Extreme Gradient Boosting, TabNet, Random Forest, and a 1D Convolutional Neural Network (CNN), achieving accuracies up to 99%, with their proposed 1D-CNN outperforming other models in terms of accuracy, sensitivity, precision, and recall. Similarly, Kunekar et al. [3] investigated conventional ML and deep learning (DL) approaches, demonstrating that Long Short-Term Memory (LSTM) networks provided superior results for seizure detection tasks.

Vieira et al. [4] proposed an LSTM-based methodology that achieved 97% validation accuracy while also focusing on feature and channel reduction. Leveraging Explainable AI (XAI), their approach attained over 95% in key metrics using only six features and five EEG channels, emphasizing the feasibility of lightweight models suitable for mobile applications.

Beyond traditional architectures, Zhu et al. [5] introduced a multidimensional Transformer fused with recurrent networks (LSTM-GRU) for seizure classification, addressing the challenge of the non-stationary and complex nature of EEG signals. Disli et al. [7] proposed a Continuous Wavelet Transform (CWT)-based depthwise CNN. Their innovative method converted multi-channel EEG data into image-like representations, achieving nearly 96% accuracy and outperforming prior methods without requiring separate feature extraction or additional classifiers.

The potential of lightweight models and TinyML for real-time applications has also been explored. Tsanika et al. [8] developed a TinyML model using iEEG data, achieving up to 99% test accuracy. Their work underscores the suitability of resource-constrained ML implementations for integration into closed-loop neurostimulation devices.

Fractal features and non-linear dynamics have been explored by Abhisek et al. [9], who employed parameters like Higuchi’s and Katz’s fractal dimensions alongside spectral features. Their method achieved remarkable performance—100% accuracy in classifying focal versus non-focal EEG signals in the Bern-Barcelona dataset, and significant improvements over state-of-the-art methods in detecting interictal and preictal states.

Brari et al. [10] proposed a novel correlation dimension-based feature extraction technique, reporting near-perfect classification accuracy on benchmark datasets while maintaining computational simplicity. Rab et al. [11] combined deep learning with domain knowledge (frequency bands, electrode locations, and temporal patterns) and introduced XAI4EEG, a framework that integrates SHAP-based explanations to improve interpretability and reduce clinicians’ validation time during seizure analysis.

Architectural innovations combining CNNs and recurrent networks have shown high effectiveness. Mallick et al. [12] designed a hybrid model incorporating 1D convolutional layers, bidirectional LSTM, GRU, and pooling layers, achieving up to 100% accuracy for binary seizure detection on the Bonn dataset and robust performance in multi-class classification tasks.

Esmaeilpour et al. (2023) [13] focused on detecting preictal states using convolutional neural networks for feature extraction, followed by ensemble classifiers like random forests and support vector machines, achieving 90.76% sensitivity on the CHB-MIT dataset. Finally, privacy-preserving methods have emerged as critical in healthcare. Suryakala et al. (2023) [14] proposed a Federated Machine Learning (FML) approach that trains models on decentralized EEG data, achieving high sensitivity (98.24%) and specificity (99.23%) while maintaining patient data confidentiality.

This study by Ahmad et al, (2024) addresses the critical challenge of early epileptic seizure prediction (ESP) by developing a patient-specific framework that leverages both handcrafted and deep learning features extracted from EEG signals. It employs a Bidirectional Long Short-Term Memory (BiLSTM) network combined with an attention mechanism to capture temporal dependencies while reducing feature dimensionality. Integrating Consumer Internet of Things (CIoT) technology enables real-time seizure prediction and alerting through cloud-based platforms, enhancing timely intervention. The model demonstrates robust performance using Leave-One-Out cross-validation, achieving high accuracy (91.39%), sensitivity (91.30%), specificity (90.06%), and a low false positive rate (0.12), highlighting its potential for practical clinical applications in smart healthcare systems.

This research [22] by Ahmad et al, (2024) proposes an Advanced Multi-View Deep Feature Learning (AMV-DFL) framework to improve epileptic seizure detection from EEG signals by combining traditional frequency and time domain features with deep features extracted via 1D convolutional neural networks. Using a multi-view forest classifier and explainable AI techniques, the method enhances interpretability and classification performance. Experimental results show that AMV-DFL outperforms traditional and state-of-the-art models by 3–4% in accuracy, supporting clinicians in better identifying key EEG features and advancing epilepsy diagnosis.

Collectively, these works illustrate the breadth of methodologies—ranging from conventional ML, deep architectures, feature engineering, explainable AI, to privacy-preserving techniques—that continue to enhance the reliability and practicality of EEG-based epileptic seizure detection systems.

Numerous studies have been conducted in the past on machine learning-based seizure detection; nevertheless, further research on diverse occurrences is necessary and essential to progress in the fields of epilepsy and seizure early detection. Through the creation and advancement of Internet of Medical Things technology, assistive strategies are being developed to make the lives of sufferers simpler. The evaluation of the ML-based framework on only two classes and all four classes was done on the same dataset in earlier works [5], which lagged behind the creation of detection and application frameworks based on three classes. With the dataset [15] we are working on 3 classes events classification as well as 4-classes events classification with various ML models development. Finally, IoMT (Internet of Medical Things) approach provide a comprehensive framework to automate patient room or environmental condition as well as triggering alert notification to caretaker under cases of emergencies.

## Proposed ML modeling approach

### Dataset

A dataset from the UCI repository source is collected, and UMAP, FFT were applied [15, 21], and details of the dataset are given in table 1 and table 2.

**Table 1:**
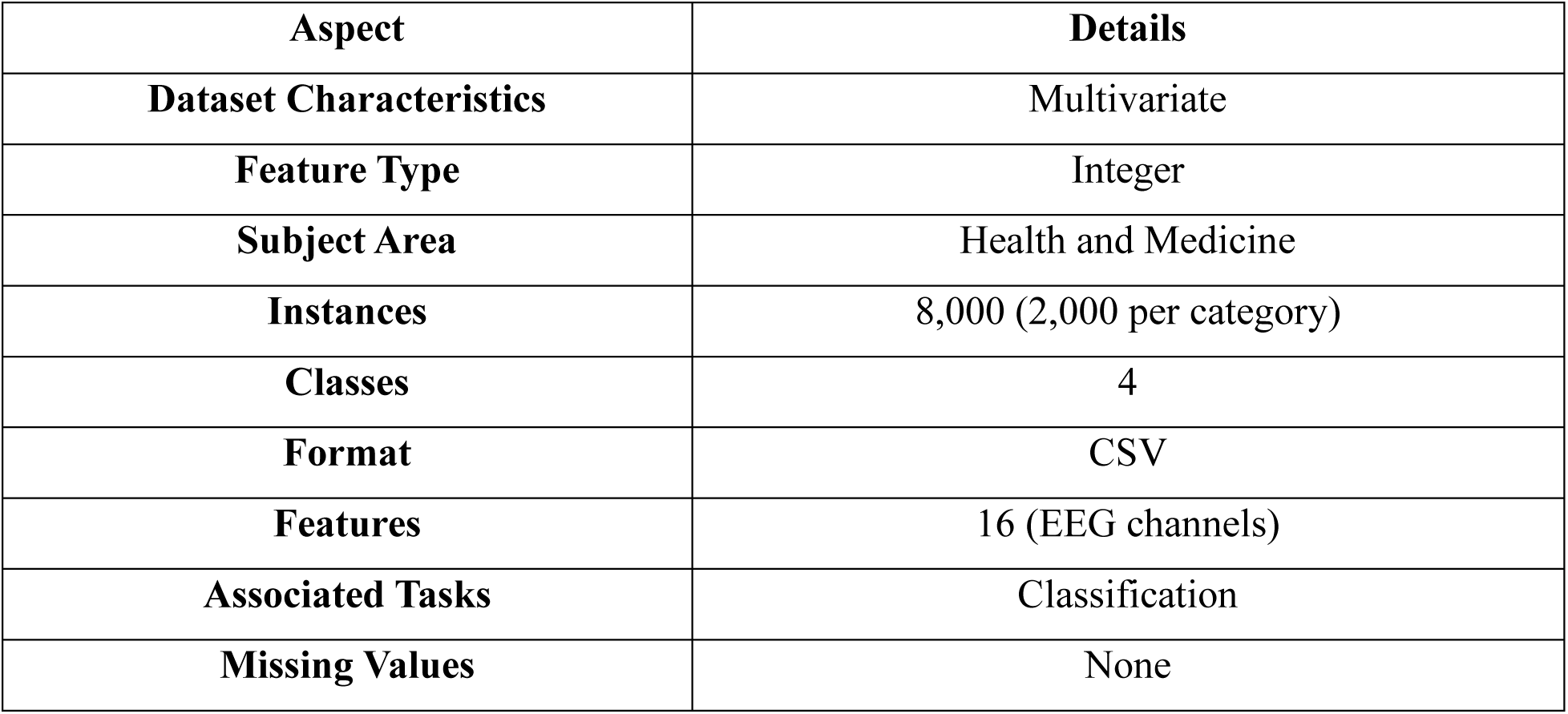
Dataset detail [15].

| Aspect | Details |
| --- | --- |
| Dataset Characteristics | Multivariate |
| Feature Type | Integer |
| Subject Area | Health and Medicine |
| Instances | 8,000 (2,000 per category) |
| Classes | 4 |
| Format | CSV |
| Features | 16 (EEG channels) |
| Associated Tasks | Classification |
| Missing Values | None |

**Table 2:**
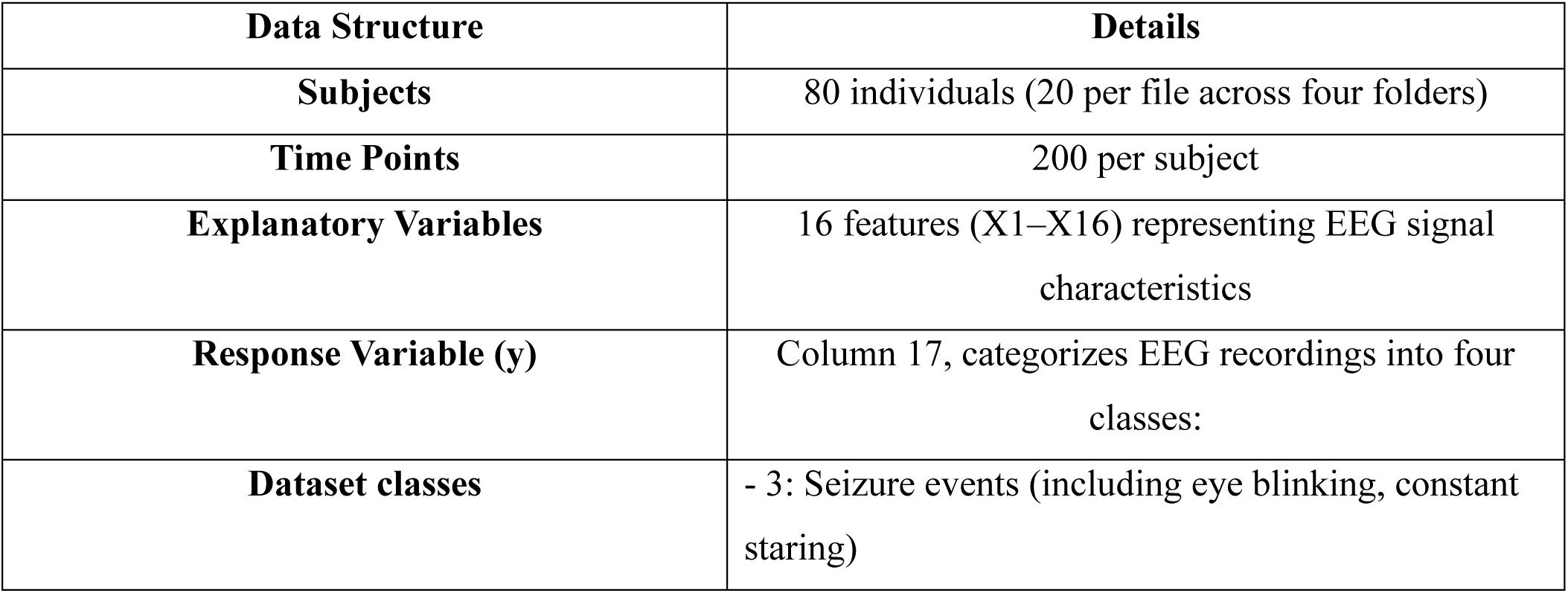
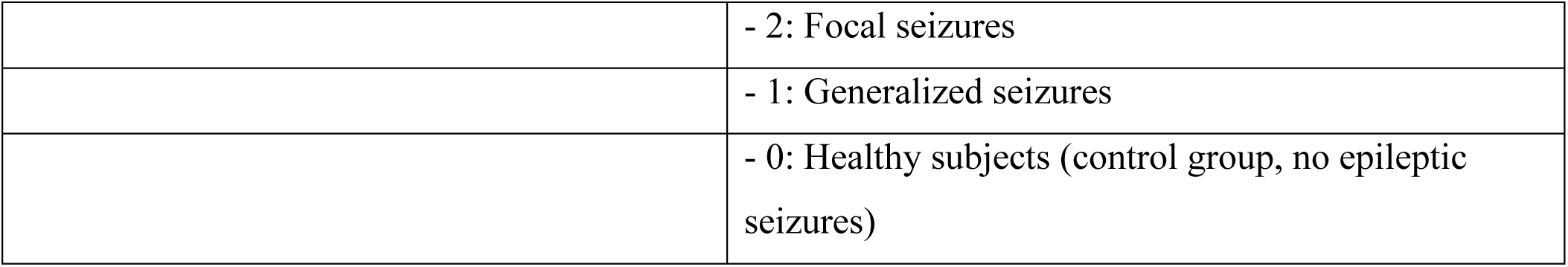
Dataset breakdown of detail.

## Preprocessing

### UMAP and FFT application

In this workflow, data preprocessing consists of two important feature transformation techniques applied one after the other to improve the raw input data prior to modeling. To begin with, the features are normalized using standard scaling, which guarantees a mean of zero and unit variance; this step aids the performance of algorithms such as random forest and UMAP by stopping features with larger scales from taking control. Next, UMAP (Uniform Manifold Approximation and Projection) is utilized as a dimensionality reduction method that captures the intrinsic temporal or structural relationships within the data, condensing the original high-dimensional feature space into a smaller set of meaningful components.

### UMAP (Uniform Manifold Approximation and Projection)

UMAP is a dimensionality reduction technique based on manifold learning and topological data analysis. Its goal is to find a low-dimensional representation of data that preserves the high-dimensional data’s essential topological structure.

### Constructing the Fuzzy Simplicial Set

UMAP begins by computing a weighted local connectivity for each point *x*_*i*_ to its neighbors *x*_*j*_. This connectivity is defined using a smooth exponential kernel:

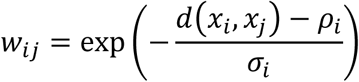

where:

- *d*(*x*_*i*_, *x*_*j*_) is the distance between points *i* and *j*.
- ρ_*i*_ is the distance to the closest neighbor, which serves as a local connectivity adjustment.
- σ_*i*_ is a scaling parameter chosen to balance local connectivity.

### Cross-Entropy Loss to Optimize Low-Dimensional Embedding

UMAP minimizes the cross-entropy between fuzzy sets in the high-dimensional and low-dimensional spaces. The loss function is given by:

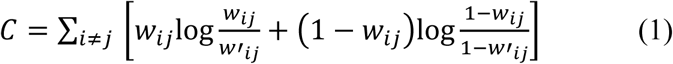

Here, *w*′_*ij*_ is the similarity between the low-dimensional embeddings *y*_*i*_ and *y*_*j*_. This is typically modeled with the following equation:

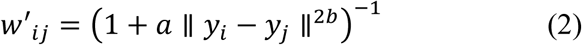

The parameters *a* and *b* control the shape of the curve.

### FFT (Fast Fourier Transform)

The FFT is an efficient algorithm for computing the Discrete Fourier Transform (DFT). It transforms a discrete time-domain signal *x*[*n*] into its frequency components *X*[*k*].

### Discrete Fourier Transform Equation

The DFT is defined as:

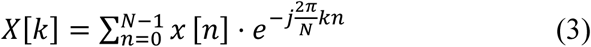

for *k* = 0,1, …, *N* − 1. In this equation:

- *x*[*n*] is the input signal at sample *n*.
- *N* is the total number of samples.
- *j* = √−1 is the imaginary unit.

The magnitude spectrum, which is often used as a feature, is calculated as:

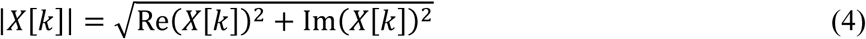

These equations highlight the core principles of how UMAP models local relationships and optimizes embeddings, and how FFT decomposes signals into their frequency components. Both methods enable the creation of richer feature representations for machine learning models.

After UMAP, the data is processed using FFT (Fast Fourier Transform) to extract spectral features by converting the scaled data into the frequency domain. This transformation emphasizes periodic patterns and frequency-based characteristics that may be essential for classification. The outputs from UMAP and FFT are concatenated to create a combined feature set that captures both temporal structure and spectral information. This provides a richer representation of the data for the Random Forest model during training and evaluation.

The figure 1 depiction shows unprocessed EEG signals from 16 channels, demonstrating temporal variations in electrical brain activity as documented in the Bangalore EEG Epilepsy dataset. Each subplot represents a particular brain region, where the horizontal axis indicates the progression of time (sample index) and the vertical axis displays voltage measured in microvolts. In the beginning (samples 0–2500), the majority of channels display high-frequency, low-amplitude activity that is characteristic of normal or inter-ictal brain states. Around sample 2500, a significant change takes place—multiple channels (especially X1, X2, X4, X5, X9, X12, and X13) exhibit a sudden increase in amplitude and a shift to rhythmic, lower-frequency oscillations, signaling the onset of a seizure (ictal state).

**Fig 1:**
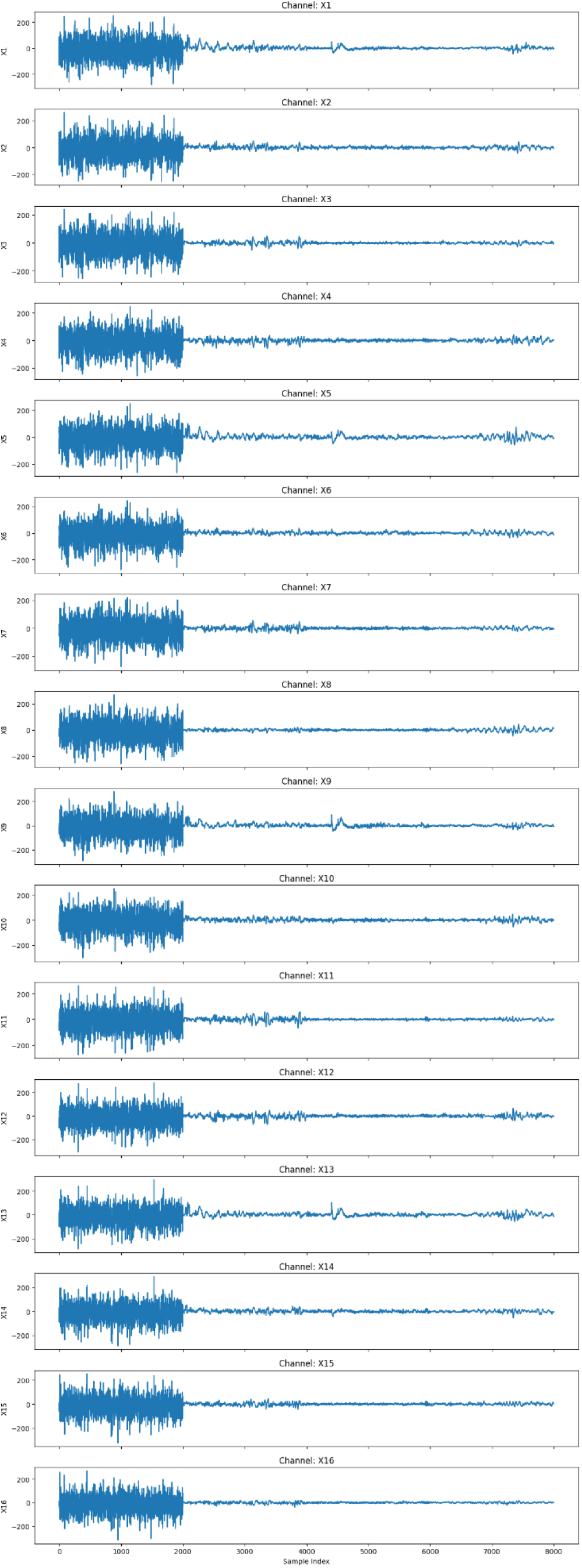
EEG signal plot for various channels.

This is followed by a post-ictal phase, where the signal flattens or reduces significantly in amplitude. The plot effectively highlights the temporal dynamics of a seizure and suggests spatial localization based on differential channel responses.

#### i. Random Forest

Random Forest is a type of ensemble learning algorithm that is mainly utilized for classification and regression tasks. During training, it constructs several decision trees and merges their predictions to enhance accuracy and manage overfitting. For classification, a majority vote is taken across all trees, while for regression, the predictions are averaged. Each tree is trained on a random data subset (bagging) and uses a random feature subset for node splitting, which adds diversity and decreases correlation among trees

#### ii. EEGNet

EEGNet is a compact convolutional neural network architecture specifically designed for EEG signal processing. It efficiently extracts spatial and temporal features from EEG data using depthwise and separable convolutions, making it well-suited for brain-computer interface (BCI) applications with limited training data and low computational cost.

#### iii. SVC

Support Vector Classification (SVC) is an algorithm for supervised machine learning that serves classification purposes. The concept is rooted in Support Vector Machine (SVM) methodology, which seeks to identify the optimal hyperplane that most effectively divides data points of varying classes within a high-dimensional space. SVC operates by maximizing the margin between the closest points (support vectors) of each class, making it suitable for both linear and non-linear classification challenges.

### Proposed Workflow

First, models are feature extracted as preprocessing was carried out with UMAP and FFT procedure and they are trained and evaluated with various evaluation metrics. In order to find out the best performance, the nested cross-validation technique is applied. This enhanced the model development as well as ensured the model’s performance isn’t compromised on different folds of the dataset. With this cross-validation technique, model performance on the entire dataset can be rigorously tested. Then, the trained model is proposed to be deployed on the proposed IoMT system-based concept to aid patients.

The various ML models are trained to make seizure detection models. A stacking ensemble models are conceptualized that performed well beyond EEGNet a common model in ML field. The figure 3 shows overall modeling concept.

**Fig 2:**
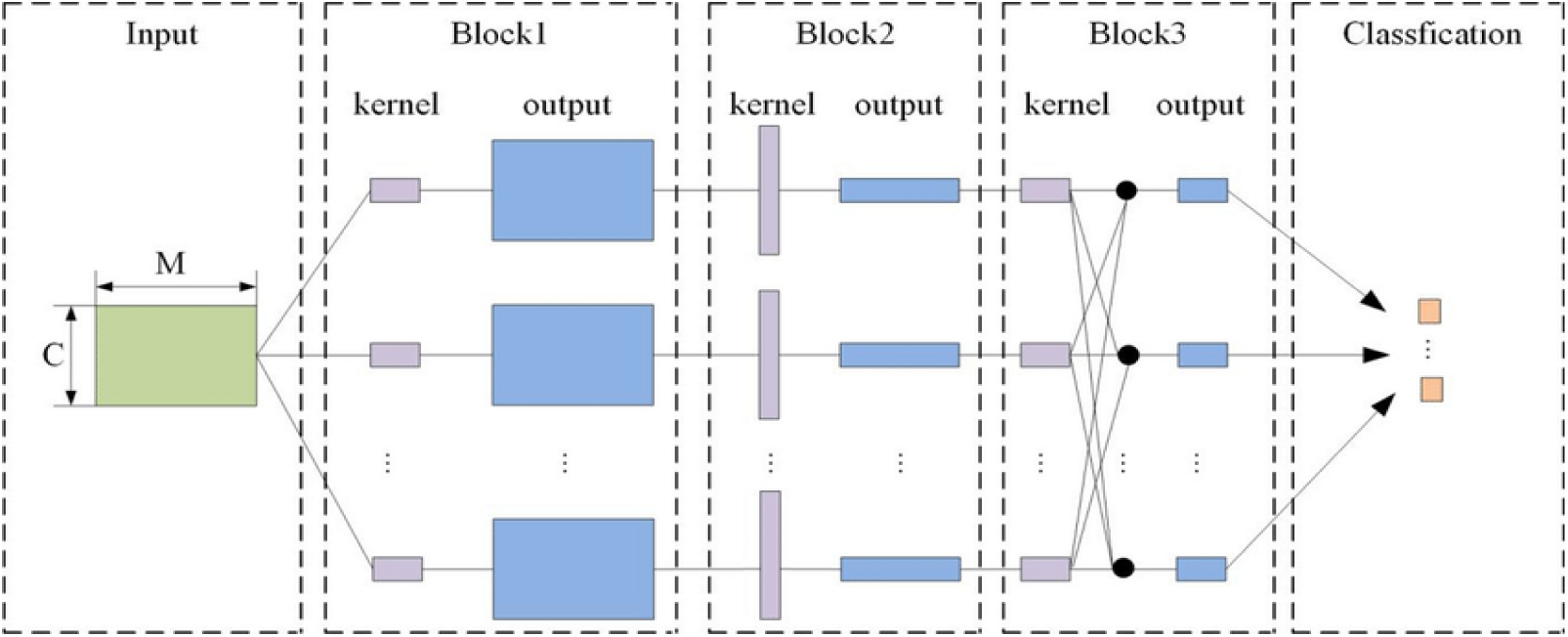
EEGNet architecture [16].

**Fig 3:**
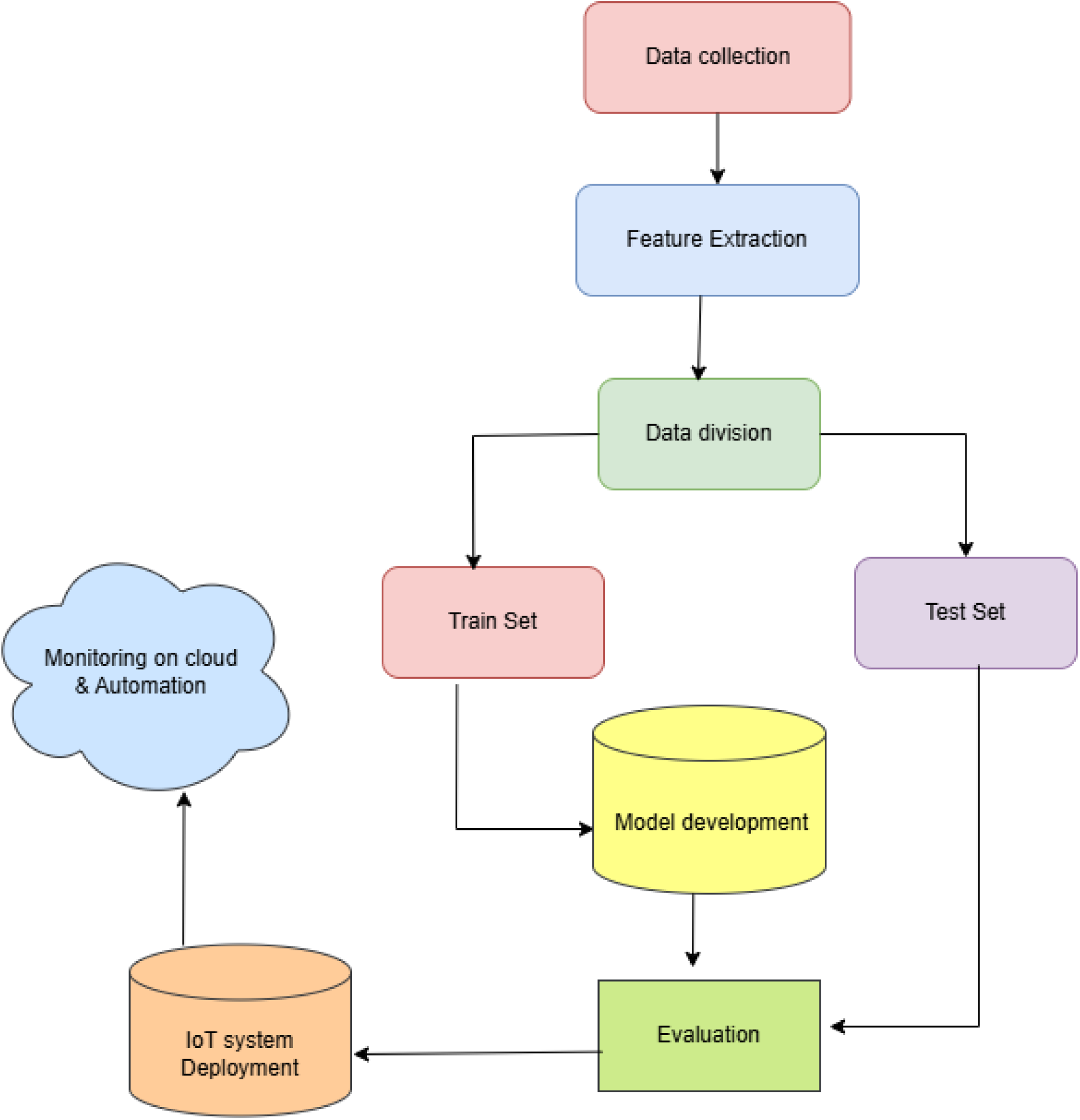
Proposed modeling concept.

### Stacking ensemble model

This model implements a two-level stacking ensemble for EEG epilepsy classification:

- Level 1: Base Learners

The base models are individually trained classifiers:

i. Random Forest: An ensemble of decision trees that uses bootstrap sampling and feature randomness to improve accuracy and control overfitting.
ii. Extra Trees: Similar to Random Forest but builds trees by selecting splits randomly, which often increases diversity among trees and reduces variance.
iii. XGBoost: A gradient boosting method that builds trees sequentially to minimize classification errors, highly effective for structured/tabular data.

- Level 2: Meta Learner

The outputs (predictions or probabilities) from the base learners are fed into a CatBoost classifier, which acts as the meta-model. CatBoost is an efficient gradient boosting algorithm known for handling categorical variables and avoiding overfitting. It learns to optimally combine the base learners’ predictions to improve the overall final accuracy.

The ensemble model as shown in figure 4 architectural diagram benefits from the diversity and complementary strengths of base learners, while the meta learner integrates these predictions for better robustness and generalization on the test data.

**Fig 4:**
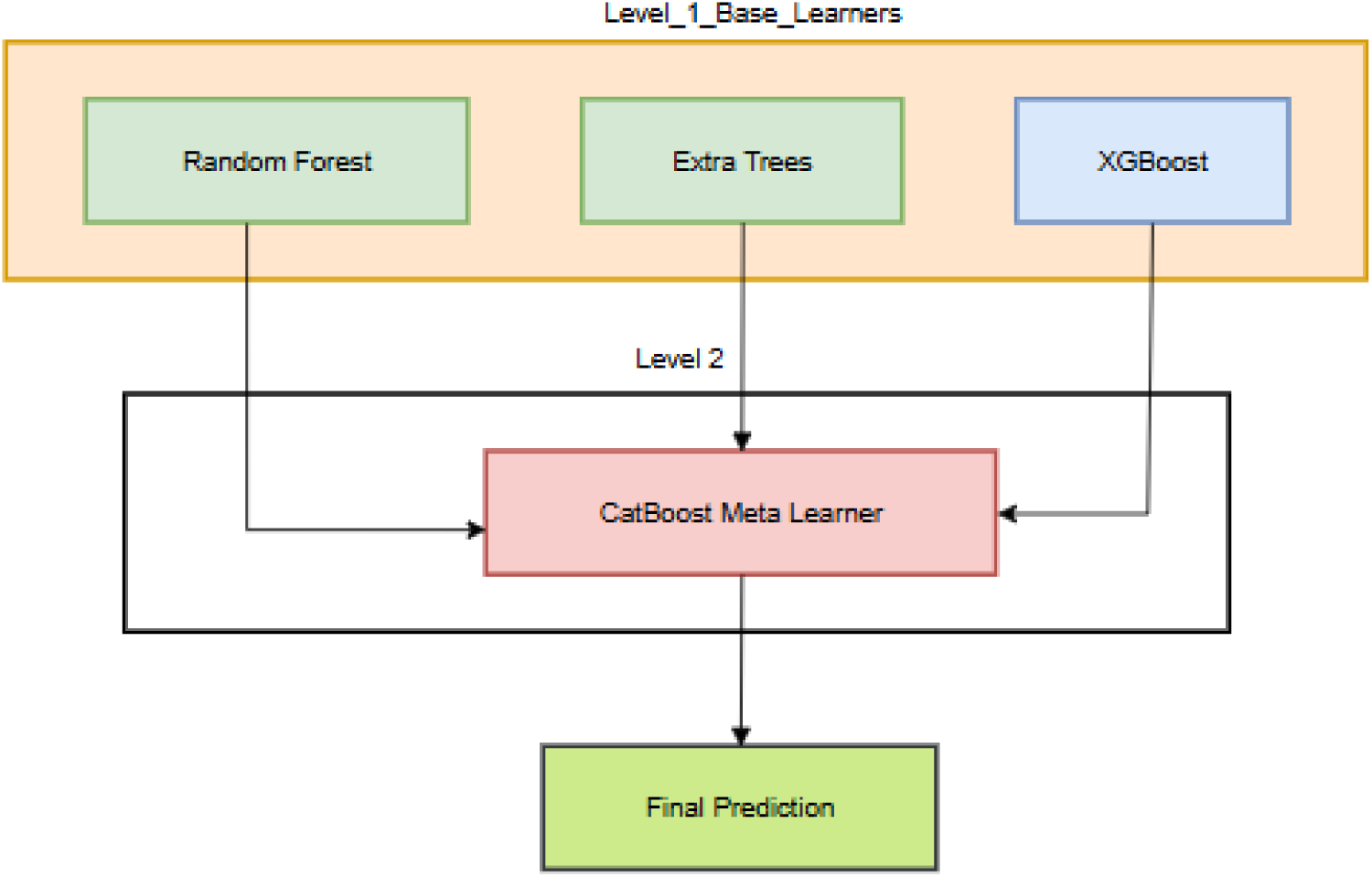
Ensemble model network.

## Evaluation metrics

Accuracy, precision, recall and F1-score: Accuracy measures the overall correctness of a classification model by calculating the proportion of total predictions that are correct. Precision evaluates how many of the positive predictions made by the model are actually correct, reflecting the model’s ability to avoid false positives. Recall (also known as sensitivity) measures the model’s ability to identify all actual positive cases by finding the proportion of true positives detected out of all real positives, thus indicating how well the model prevents false negatives. The F1-score is the harmonic mean of precision and recall, providing a balanced metric that combines both aspects to obtain one performance measure, especially useful when the class distribution is imbalanced [17].

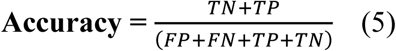

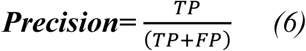

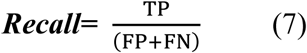

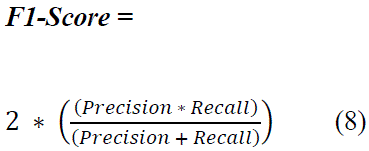

## • LIME analysis

LIME (Local Interpretable Model-agnostic Explanations) is an interpretability technique used to explain the predictions of any black-box machine learning model. It performs operations by perturbing the input data locally around a prediction and fitting a simple, interpretable model— like SVC or other models—on this perturbed dataset to approximate the behavior of the complex model in that local region. This helps the users to understand which features contributed most to a specific prediction, helping build trust and transparency in model decisions without further need to understand the entire model’s architecture.

## Results and Discussions

The neural network models like EEGNet suffered overfitting, while the two-level ensemble model with approximately 97% accuracy depicted a strong outcome. The models went through cross-validation evaluation in order to evaluate performances across the entire dataset’s different folds.

## Model wise performances

EEGNet: The EEGNet model achieved training and testing accuracies of 69.65% and 69.63%, respectively. There are indications that the model has not learned adequately, and although regularization and dropouts were subsequently implemented, there was no enhancement in the model’s performance. The primary problem with this approach was that the neural networks could not produce stable outcomes and experienced significant overfitting, despite the application of regularization and other methods to mitigate these issues.

The figure 5 shows the “Average FFT Spectrum by Class” that suggests a frequency-domain analysis of EEG signals, grouped by distinct classes (e.g., different brain states or tasks). The “EEGNet Accuracy” and “Train validation” sections indicate the model’s performance during training, with metrics tracked across epochs (0 to 7). The inclusion of UMAP implies dimensionality reduction to highlight patterns or separability in the temporal features, correlating with the model’s accuracy. Overall, the figure combines signal processing (FFT), machine learning (EEGNet), and visualization (UMAP) to analyze and interpret EEG data.

**Fig 5:**
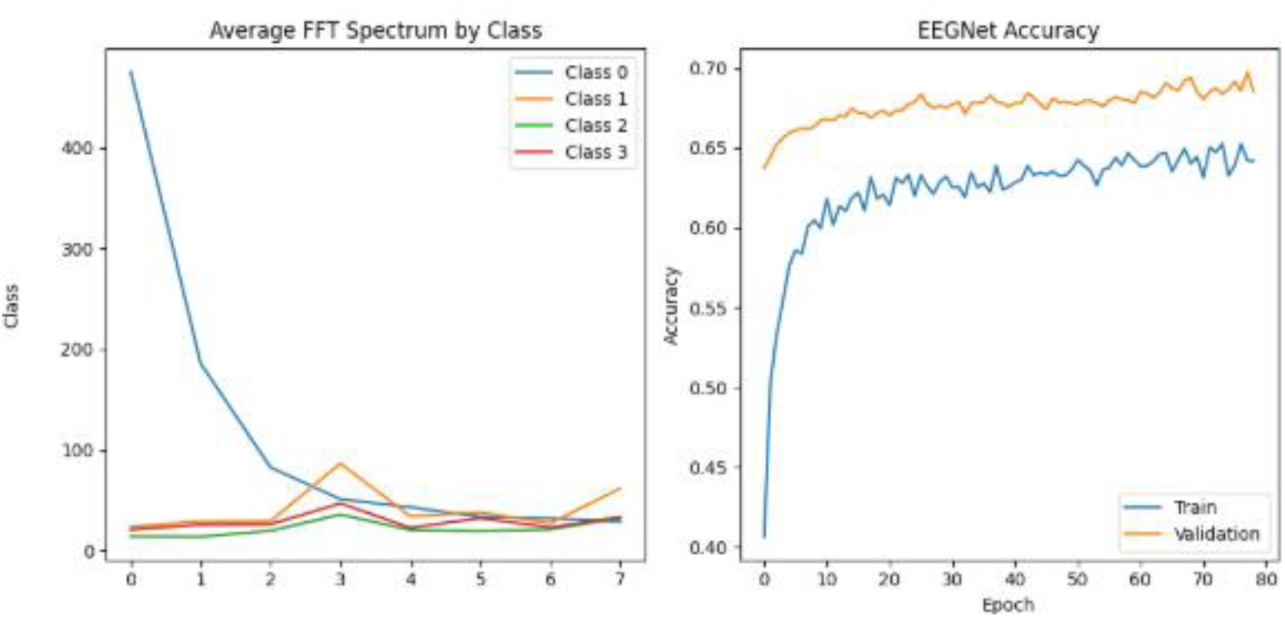
Model history plots.

## Random Forest classifier

The model random forest obtained accuracy of 94.19% and training accuracy of 100%. The model was cross validated across 10-folds in table 3 to check for any signs of overfitting.

**Table 3:** Nested Cross-Validation Results.

| Fold | Test Accuracy |
| --- | --- |
| Fold 1 | 0.9525 |
| Fold 2 | 0.9425 |
| Fold 3 | 0.9375 |
| Fold 4 | 0.9525 |
| Fold 5 | 0.9525 |
| Fold 6 | 0.9500 |
| Fold 7 | 0.9475 |
| Fold 8 | 0.9475 |
| Fold 9 | 0.9550 |
| Fold 10 | 0.9712 |
| **Mean** | **0.9509** |
| **Std Dev** | **0.0084** |

## Ensemble Stacking classifier

The model confusion matrix shown in figure 6 shows how well the model performed and predicted on testing data. The model obtained a test accuracy of 97.06% and a training accuracy of 100%. The model ensemble stacking classifier with 399 correct predictions for class ‘0’ and 394 for class 1 and 389 correct predictions for class ‘2’ and finally ‘385’ correct predictions for class ‘3’ could successfully classify various epileptic events. The models overfitting was evaluated with nested cross fold validation for 10 folds as tabulated. The nested cross fold validation in table 4 shows 91.51% test accuracy with only 0.0162 as standard deviation showing models consistent learning over entire dataset folds.

**Fig 6:**
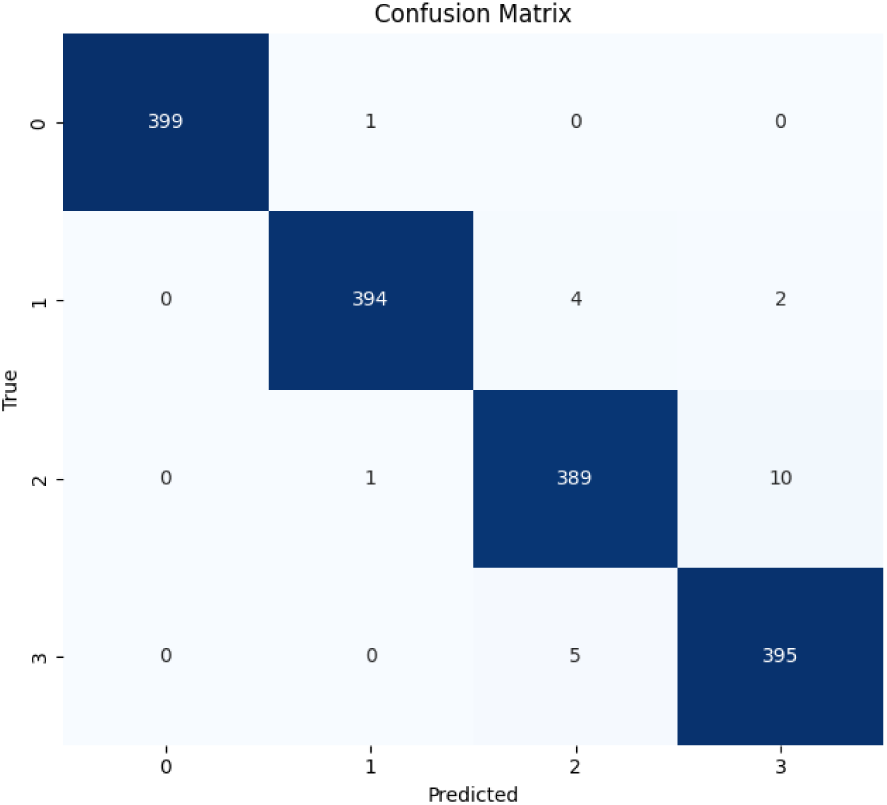
Ensemble stacking classifier model confusion matrix plot.

**Table 4:** Nested cross validation result:

| **Fold** | **Test Accuracy** |
| --- | --- |
| Fold 1 | 0.9250 |
| Fold 2 | 0.9237 |
| Fold 3 | 0.9375 |
| Fold 4 | 0.8962 |
| Fold 5 | 0.9287 |
| Fold 6 | 0.9300 |
| Fold 7 | 0.9125 |
| Fold 8 | 0.9075 |
| Fold 9 | 0.8825 |
| Fold 10 | 0.9075 |
| **Mean Accuracy** | **0.9151** |
| **Standard Deviation** | **0.0162** |

## LIME intrepretability

The LIME explanation for test instance 0 as shown in figure 7 highlights how specific feature ranges influenced the model’s prediction. Among the features, Feature_15 in the range 0.00 to 0.16 contributed most positively to the prediction with a weight of +0.1316, followed by Feature_7 between –0.18 and 0.00 (+0.0881), and Feature_12 between 0.00 and 0.23 (+0.0667). These features increased the model’s confidence in its decision. On the other hand, some features had a negative influence, decreasing the likelihood of the predicted class. Notably, Feature_2 greater than 0.18 had the strongest negative impact (–0.0616), followed by Feature_5 and Feature_4, both greater than 0.23, with weights of –0.0489 and –0.0476, respectively. Additional features such as Feature_1 > 0.25 and Feature_9 > 0.27 also slightly reduced the prediction confidence. Overall, LIME provides a local explanation by quantifying how specific feature intervals either supported or opposed the model’s final decision for this particular instance. The figure 7 and figure 8 shows LIME summary and local explanations plot. This shows feature 15 (channel 16 data) as most significant in predictions.

**Fig 7:**
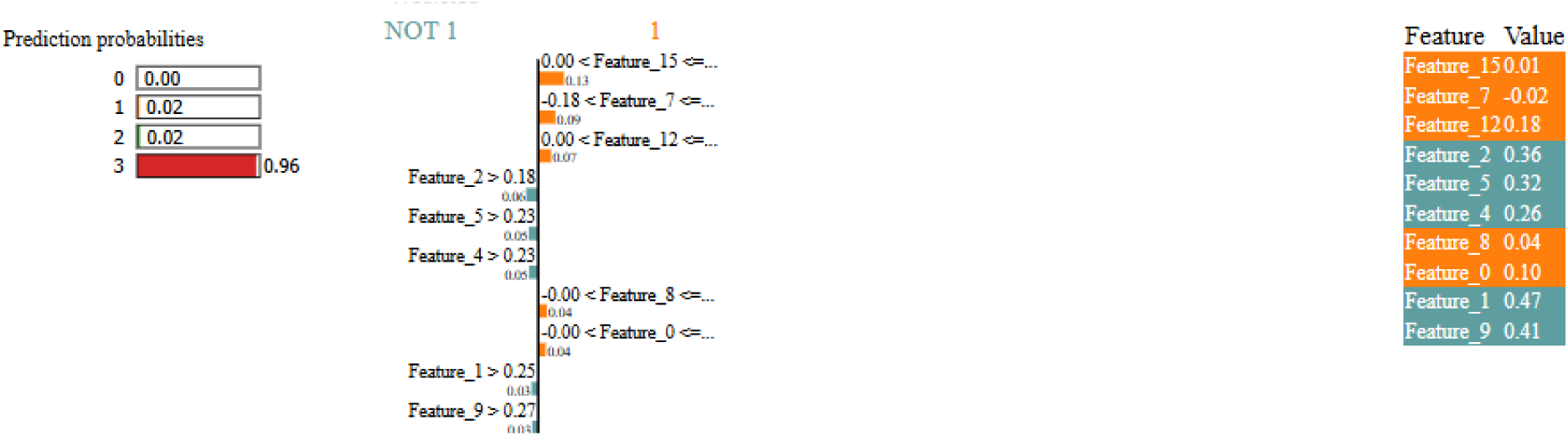
LIME analysis plot.

**Fig 8:**
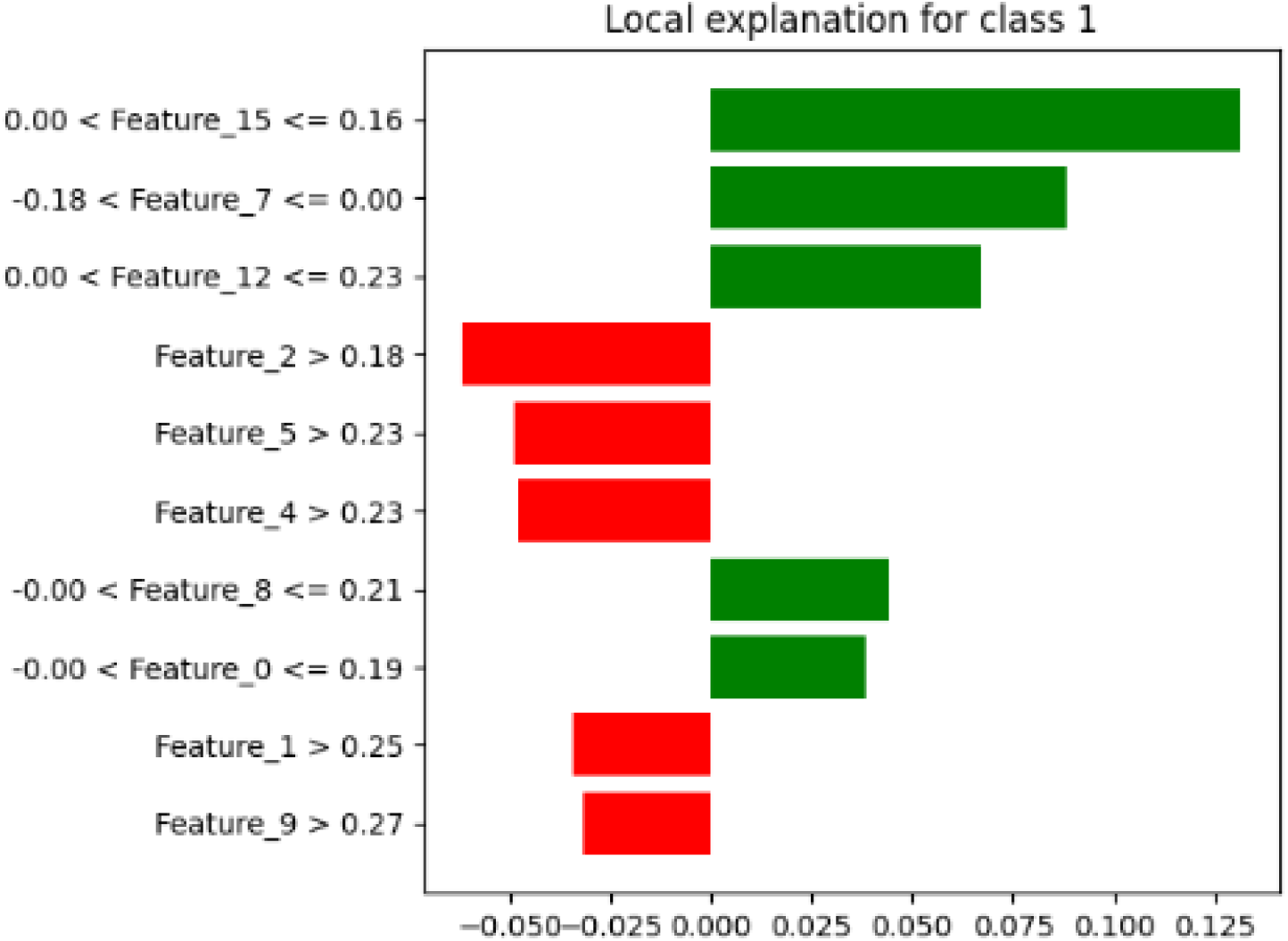
Local explanations plot.

Besides, the Support Vector Classifier model obtained a training accuracy of 66.22% test Accuracy of 65.81% and the model catboost obtained a test Accuracy: of 90.38%. The model hyperparametrs are given in table 5 and table 6 shows the summarized classification report for all models utilized.

**Table 5:**
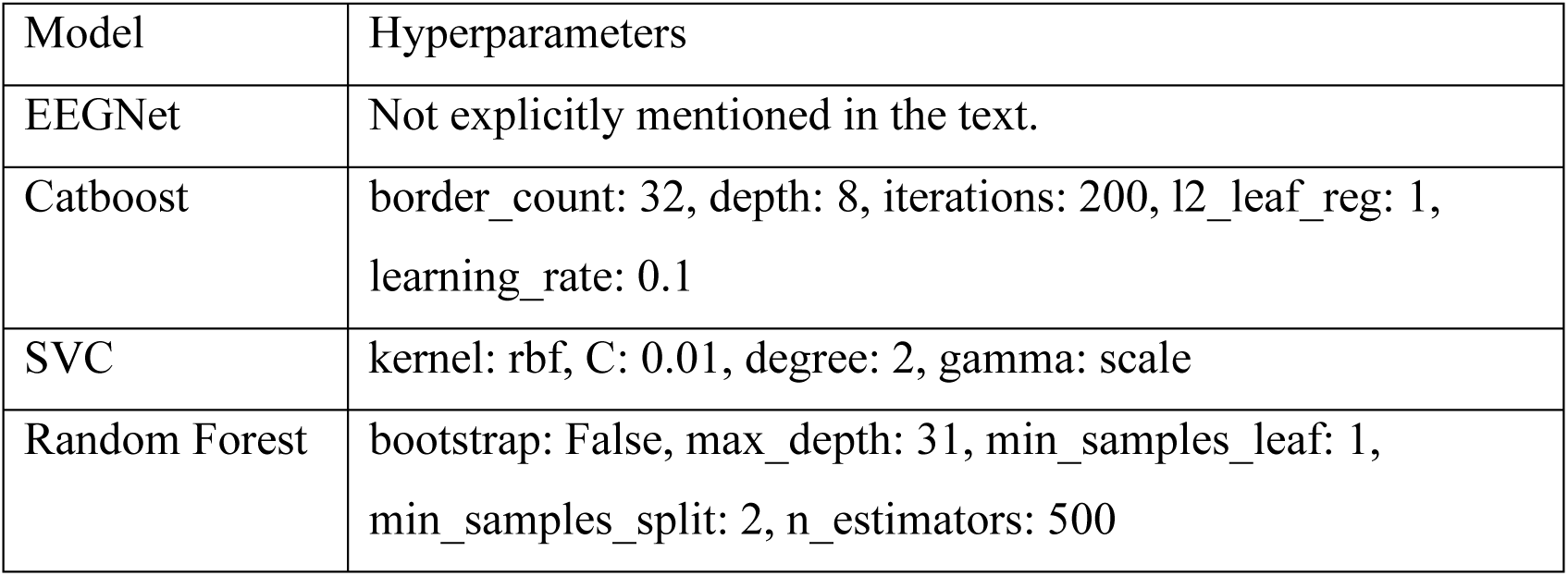
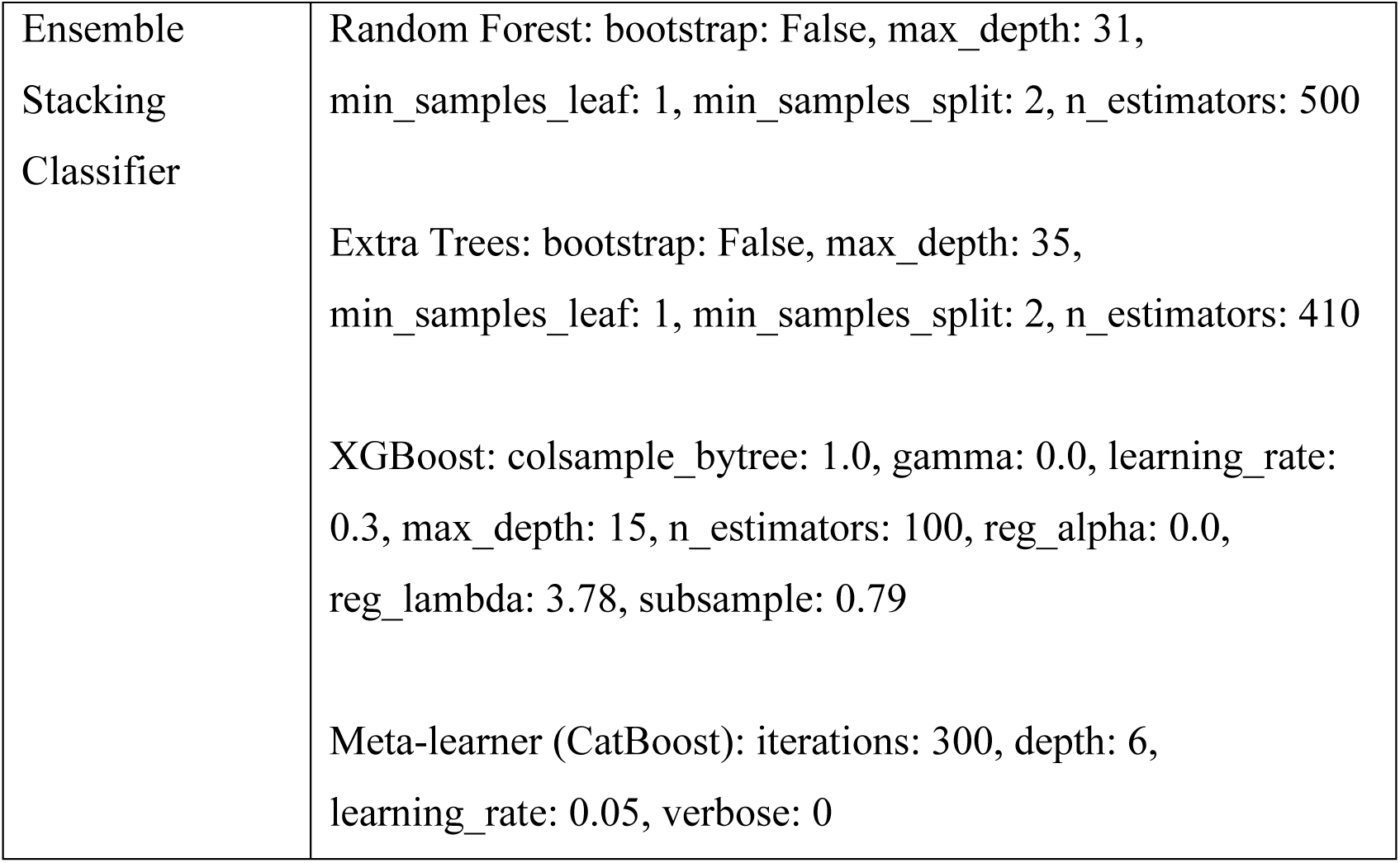
Model wise hyperparamters.

| Model | Hyperparameters |
| --- | --- |
| EEGNet | Not explicitly mentioned in the text. |
| Catboost | border_count: 32, depth: 8, iterations: 200, l2_leaf_reg: 1, learning_rate: 0.1 |
| SVC | kernel: rbf, C: 0.01, degree: 2, gamma: scale |
| Random Forest | bootstrap: False, max_depth: 31, min_samples_leaf: 1, min_samples_split: 2, n_estimators: 500 |
| Ensemble<br>Stacking<br>Classifier | Random Forest: bootstrap: False, max_depth: 31, min_samples_leaf: 1, min_samples_split: 2, n_estimators: 500<br>Extra Trees: bootstrap: False, max_depth: 35, min_samples_leaf: 1, min_samples_split: 2, n_estimators: 410<br>XGBoost: colsample_bytree: 1.0, gamma: 0.0, learning_rate: 0.3, max_depth: 15, n_estimators: 100, reg_alpha: 0.0, reg_lambda: 3.78, subsample: 0.79<br>Meta-learner (CatBoost): iterations: 300, depth: 6, learning_rate: 0.05, verbose: 0 |

**Table 6:** Classification report summary.

| Model | Class | Precision | Recall | F1-Score | Support |
| --- | --- | --- | --- | --- | --- |
| EEGNet | 0 | 1 | 1 | 1 | 400 |
|  | 1 | 0.64 | 0.76 | 0.69 | 400 |
|  | 2 | 0.6 | 0.81 | 0.69 | 400 |
|  | 3 | 0.48 | 0.21 | 0.3 | 400 |
|  | Accuracy |  |  | 0.7 | 1600 |
|  | Macro Avg | 0.68 | 0.7 | 0.67 | 1600 |
|  | Weighted Avg | 0.68 | 0.7 | 0.67 | 1600 |
| Catboost | 0 | 1 | 1 | 1 | 400 |
|  | 1 | 0.96 | 0.86 | 0.91 | 400 |
|  | 2 | 0.81 | 0.89 | 0.84 | 400 |
|  | 3 | 0.86 | 0.87 | 0.86 | 400 |
|  | Accuracy |  |  | 0.9 | 1600 |
|  | Macro Avg | 0.91 | 0.9 | 0.9 | 1600 |
|  | Weighted Avg | 0.91 | 0.9 | 0.9 | 1600 |
| SVC | 0 | 0.98 | 0.99 | 0.98 | 400 |
|  | 1 | 0.61 | 0.68 | 0.64 | 400 |
|  | 2 | 0.54 | 0.8 | 0.65 | 400 |
|  | 3 | 0.41 | 0.15 | 0.22 | 400 |
|  | Accuracy |  |  | 0.66 | 1600 |
|  | Macro Avg | 0.63 | 0.66 | 0.62 | 1600 |
|  | Weighted Avg | 0.63 | 0.66 | 0.62 | 1600 |
| Random Forest | 0 | 1 | 0.99 | 1 | 400 |
|  | 1 | 0.95 | 0.93 | 0.94 | 400 |
|  | 2 | 0.88 | 0.96 | 0.92 | 400 |
|  | 3 | 0.94 | 0.88 | 0.91 | 400 |
|  | Accuracy |  |  | 0.94 | 1600 |
|  | Macro Avg | 0.94 | 0.94 | 0.94 | 1600 |
|  | Weighted Avg | 0.94 | 0.94 | 0.94 | 1600 |
| Ensemble Stacking | 0 | 1 | 1 | 1 | 400 |
| Classifier | 1 | 0.97 | 0.96 | 0.96 | 400 |
|  | 2 | 0.95 | 0.96 | 0.96 | 400 |
|  | 3 | 0.97 | 0.96 | 0.96 | 400 |
|  | Accuracy |  |  | 0.97 | 1600 |
|  | Macro Avg | 0.97 | 0.97 | 0.97 | 1600 |
|  | Weighted Avg | 0.97 | 0.97 | 0.97 | 1600 |

**Table 7:** Comparison to state of the art approaches.

| **Ref** | **Dataset(s) Used** | **Techniques / Models Applied** | **Cross-Validation Used** | **Reported Accuracy (%)** |
| --- | --- | --- | --- | --- |
| [19] | BONN (UCI),<br>BEED | UMAP + FFT,<br>SeqBoostNet (Stacking of<br>ML + DL models) | No | 96.71 (BEED),<br>95.91<br>(Focal/Gen) |
| [20] | BONN (UCI),<br>CHB-MIT<br>(PhysioNet), BEED | AIFFT-NS (adaptive FFT)<br>+ Improved Deep Belief<br>Network (IDBN) | No | 96.16 (BEED) |
| **This Work** | BEED | UMAP + FFT + Ensemble<br>Stacking (TLE Network),<br>nested cross-validation | Yes (Nested<br>10-fold) | 97.06 (BEED) |

The performance of the EEGNet and SVC models is similar and hasn’t shown better outcome compared to ensemble and boosting models. The two models reached an overall accuracy of approximately 66–70%. They excelled in class 0 (achieving flawless precision, recall, and F1-score), but their performance declined markedly for the remaining classes. In particular, the precision and recall scores for classes 1, 2, and 3 are significantly lower, suggesting that the models find it difficult to accurately recognize these classes, especially class 3, which poses the greatest challenge for both models. This imbalance is illustrated by the macro and weighted averages, which show overall scores of approximately 0.62–0.67. The comparison to state of the art approaches is shown in table 6. Which shows that model has been successful to demonstrate an extra ordinary outcome performing better than state of the art models on same dataset.

## Limitations and Future works

Thus, the proposed approach shows a significant concept in EEG signal classification for detecting various kinds of seizures and is vital to identify their impacting features, the channels that much affect the prediction, as well as choosing the best model for performance enhancement. With current work, the model is applicable for realtime edge and IoT devices integrations. As proposed work has depicted an IoMT concept for aiding patients, a real-time evaluation couldn’t be made due to a lack of EEG headsets and more advanced devices. In the future, rigorous testing for such prototyping can be carried out. The model can be employed with various techniques for improvement of neural network models and algorithms development.

## Conclusion

To conclude, an ML model like stacking ensemble architecture has a higher potential to gather greater performance for EEG results. The models trained on the dataset show that prior to various epileptic conditions, early warning can be triggered using ML-based detection via victim EEG data. The UMAP and FFT allowed the features preprocessing to get better. This approach can be useful for deployment in real-time prototyping for IoMT edge and medical device development. With this approach, future intelligent biomedical systems based on EEG can be enhanced.

## Data Availability

Data is cited in manuscript available on UCI repository.

https://archive.ics.uci.edu/dataset/1134/beed:+bangalore+eeg+epilepsy+dataset

